# The social media response to twice-weekly mass asymptomatic testing in England

**DOI:** 10.1101/2021.08.19.21262284

**Authors:** Amelia Dennis, Charlotte Robin, Holly Carter

## Abstract

**Background:** From 9^th^ April 2021, everyone in England has been encouraged to take two COVID-19 tests per week. This is the first time that national mass asymptomatic testing has been introduced in the UK and the effectiveness of the policy depends on uptake with testing and willingness to self-isolate following a positive test result. This paper examines attitudes towards twice-weekly testing, as well as barriers and facilitators to engaging in testing.

**Methods:** Between 5^th^ April and 28^th^ May 2021 we searched Twitter, Facebook, and online news articles with publicly available comment sections to identify comments relating to twice-weekly testing. We identified 5783 comments which were then analysed using a framework analysis.

**Results:** We identified nine main themes. Five themes related to barriers to engaging in testing: low perceived risk from COVID-19; mistrust in the government; concern about taking a test; perceived ineffectiveness of twice-weekly testing policy; and perceived negative impact of twice-weekly testing policy. Four themes related to facilitators to engaging in testing: wanting to protect others; positive perceptions of tests; a desire to return to normal; and perceived efficacy for reducing asymptomatic transmission.

**Conclusions:** Overall, the comments identified indicated predominately negative attitudes towards the twice weekly testing policy. Several recommendations can be made to improve engagement with twice weekly testing, including: 1) communicate openly and honestly about the purpose of testing; 2) provide information about the accuracy of tests; 3) provide financial support for those required to self-isolate, and; 4) emphasise accessibility of testing.

## Background

From 9^th^ April 2021, everyone in England has been encouraged to take two COVID-19 tests per week, as part of a plan to reduce COVID-19 transmission [1]. The tests used for twice-weekly testing are lateral flow antigen tests (LFTs) that can be done at home and give results in under 30 minutes. The aim of this type of large-scale asymptomatic testing is to rapidly identify individuals who are infectious with COVID-19. The success of national asymptomatic testing relies on high levels of testing and subsequent isolation for those who test positive [2, 3]

Mass asymptomatic testing has been trialled in some places in England such as in Liverpool. The testing in Liverpool led to an 18% increase in case detection (compared with control areas), with models estimating that between 850 – 6600 further cases were prevented [4]. In the first month testing uptake was low, with only 25% of residents taking part [5]. However, after six months uptake had risen to 57%, with 47% of those who took part in testing having more than one test [4]. Various barriers were identified that may have contributed to low uptake, including poor site access, concerns over queuing, and lack of trust in authorities [6]. In mass weekly testing in Southampton, high-levels of communication, trust and convenience resulted in improved engagement with testing. Those who did not get tested reported reasons such as mistrust over personal data, the potential loss of money due to a positive test, and the environmental impact of tests [7]. Additionally, in asymptomatic testing of students at the University of Nottingham, facilitators to getting tested included desire to control the virus and the experience of taking a test being quick, whereas barriers included guilt about the impact on others (e.g., others having to isolate) and the mental health impact of isolation [8].

To ensure that the new twice-weekly testing programme is effective in reducing the spread of COVID-19, it will be essential that uptake with the programme is as high as possible. It is therefore important to understand public attitudes towards regular testing, including barriers and facilitators to testing uptake. This will enable barriers to be addressed, and testing uptake to be increased.

### The current study

Previous studies have assessed attitudes, barriers and facilitators towards asymptomatic testing. However, this has been done at the local or regional level and has typically involved individuals attending a test centre; the current twice-weekly testing programme is the first attempt to carry out national mass asymptomatic testing in England. The current study will extend existing research by using rapid qualitative analysis of social media data to explore public attitudes towards national twice-weekly testing. In addition, using social and online media narratives will enable insights to be captured from people who may not usually take part in standard evaluation techniques, such as interviews or surveys. The aim of this project was to generate insights into public attitudes to twice weekly testing, and to identify barriers and facilitators to engaging in the twice weekly testing programme.

## Method

### Sampling

Data was collected through publicly accessible social and online media sources, including Twitter, Facebook, and comment sections from national newspapers. All sampling captured comments between 5^th^ April 2021 (the date twice-weekly testing was announced) and 28th May 2021, when the data were collected. We identified a total of 5783 comments: 485 comments from Twitter; 3776 comments from Facebook; and 1522 comments from newspaper articles, see Table 1 for more details on sampling.

**Table 1.** Details of Sampling and Comments

|  |  | Number of Comments |
| --- | --- | --- |
| Twitter | One Government Account | 138 |
|  | One Government Official | 347 |
|  | Total for Twitter | 485 |
| Facebook | Daily Express | 93 |
|  | Daily Mail | 662 |
|  | The Mirror | 253 |
|  | The Metro | 937 |
|  | The Independent | 119 |
|  | The Sun | 305 |
|  | The Guardian | 734 |
|  | The Telegraph | 673 |
|  | Total Facebook | 3775 |
| News Articles | Article with Most Comments | 1000 |
|  | Article with The Second Most Comments | 522 |
|  | Total News Articles | 1522 |
|  | Total Number of Comments | 5783 |

### Twitter

We searched Twitter using “twice a week test” and identified official accounts that had tweeted about the twice-weekly testing. We sampled replies to official account tweets that had over 10 replies.

### Facebook

We sampled the Facebook pages for eight national newspapers: Daily Express, Daily Mail, The Guardian, The Mirror, The Metro, The Independent, The Telegraph, and The Sun. We searched these pages using the term “twice a week tests”. Comments were collected from the article on each newspaper site which had the most comments. Therefore, the comments from eight Facebook posts were sampled.

### News Articles

We sampled comments from one national newspaper, the Daily Mail; we attempted to sample comments from other newspapers, but comment sections were either not available or had been disabled. We found articles by using the search term “twice a week tests” then we sampled the comments from the two articles with the most comments.

### Analysis

Prior to analysis, we depersonalised the data by removing any identifiable information, such as names and locations. Data were analysed using framework analysis, a thematic approach that is often used in research that has implications for policy [9]. NVivo was used to conduct the five steps of framework analysis [10]: familiarisation with the data; identifying initial codes relevant to the research; indexing broad themes; charting the data into an analytic framework; and defining and clarifying themes in relation to other themes. The lead author analysed all the data and a second coder analysed 485 (∼8%) of the data. The team then met to discuss any discrepancies and ensure consistency.

## Results

Nine main themes were identified. Five themes focused on barriers to testing that included: low perceived risk from COVID-19; mistrust in the government; concern about taking a test; perceived ineffectiveness of testing policy; and perceived negative impact of twice-weekly testing policy. The other four themes related to facilitators to testing, including: wanting to protect others; positive perceptions of the tests; a desire to return to normality; and perceived efficacy for reducing asymptomatic transmission. See Table 2 for an overview of all themes and sub-themes.

**Table 2.**
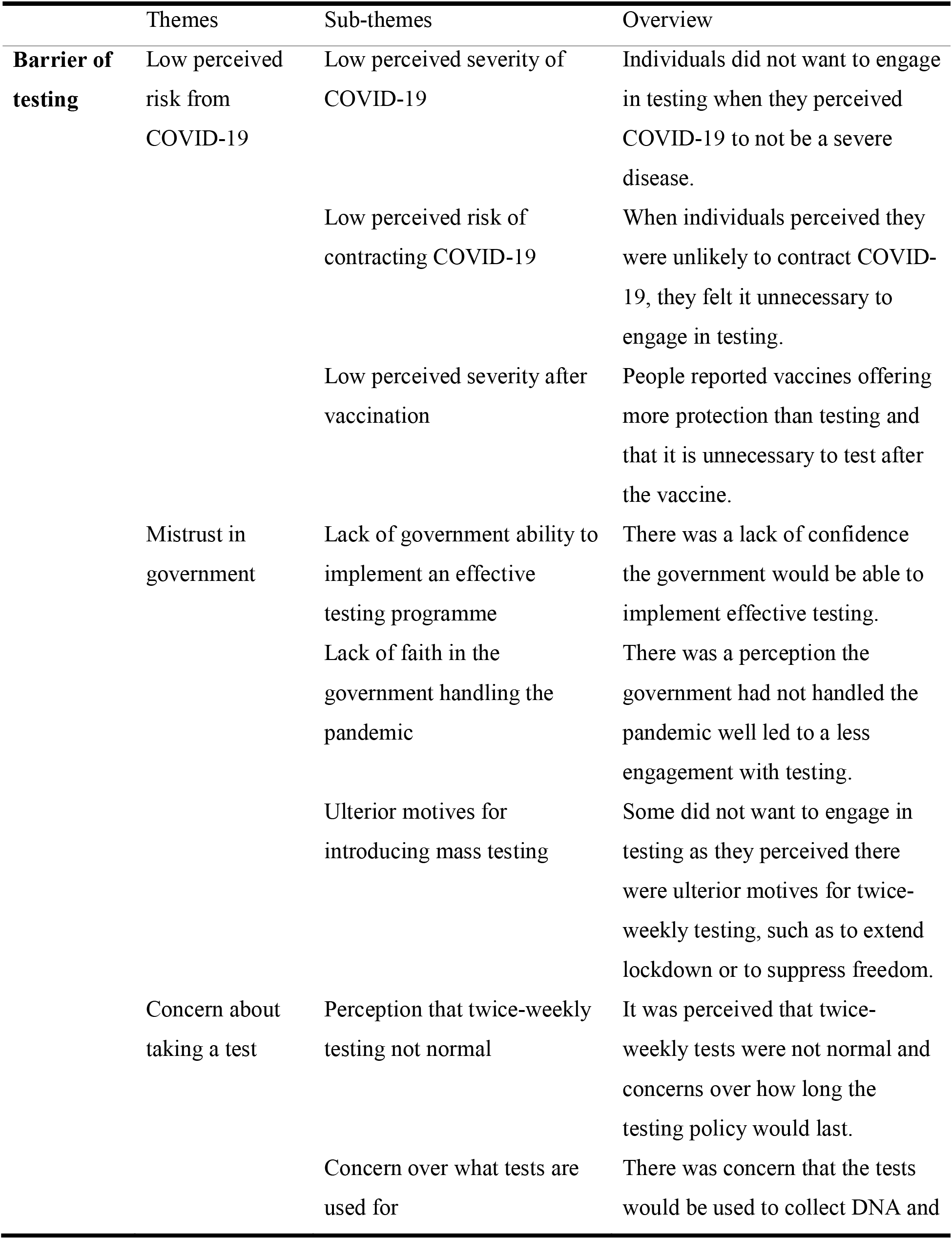
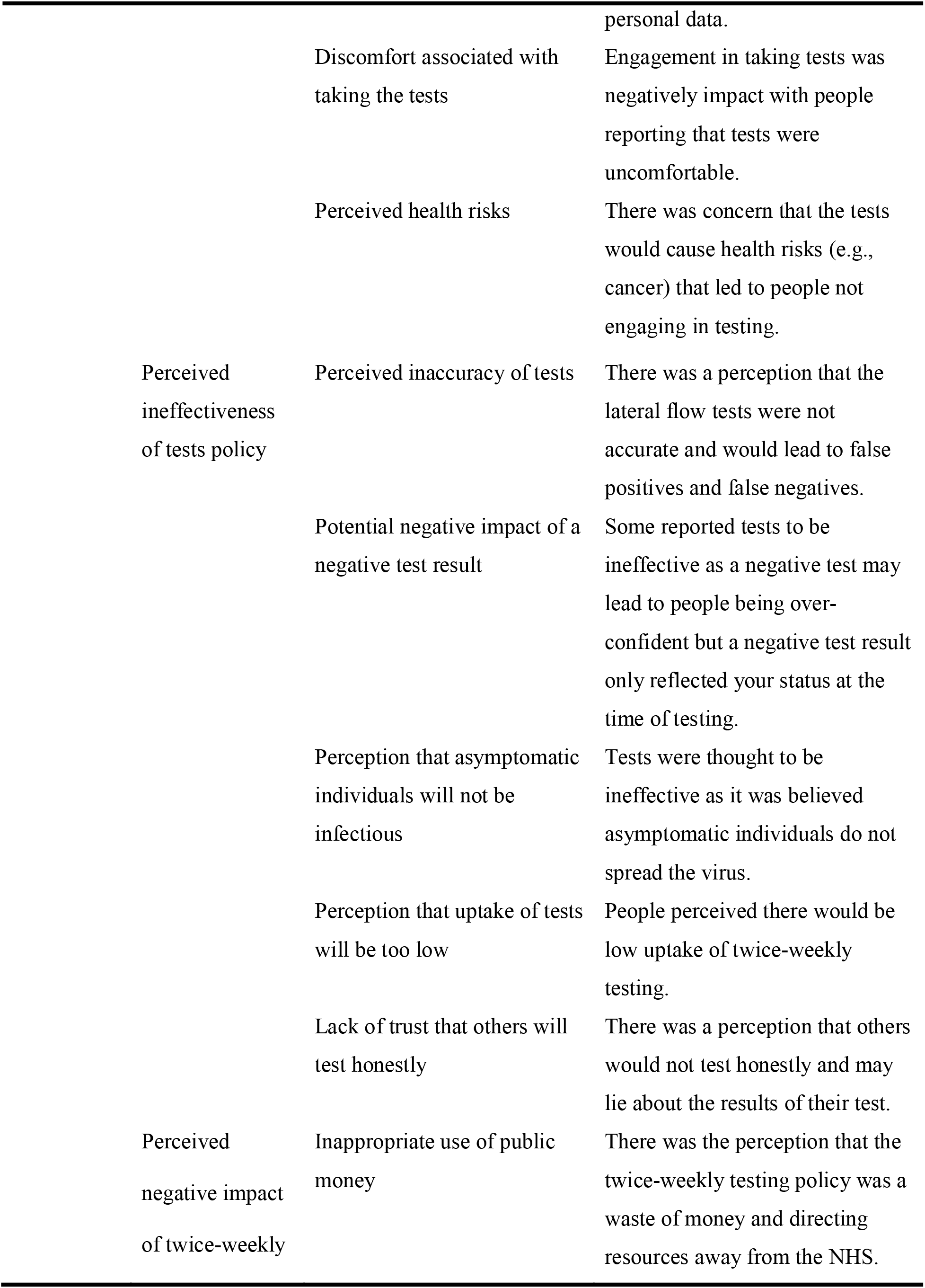
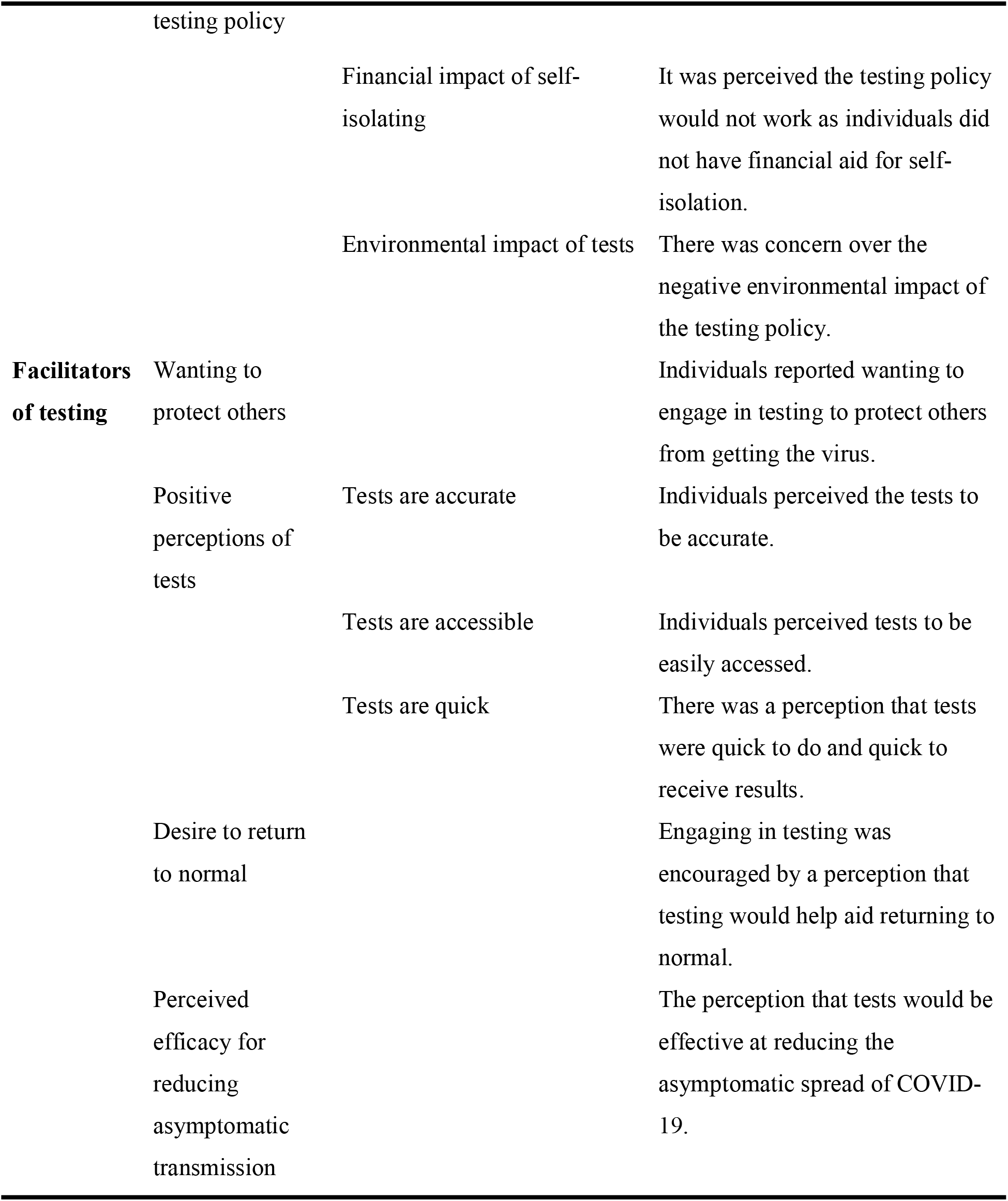
Overview of themes and sub-themes

### Low perceived risk from COVID-19

Low perceived risk from COVID-19 was a barrier to engaging in testing that revolved around three sub-themes: low perceived severity of COVID-19, low perceived risk of contracting COVID-19, and low perceived severity after vaccination.

#### Low perceived severity of COVID-19

There was the perception that COVID-19 was not a severe disease, which was largely based on the recovery rate: “*you mean the data that says the virus has a 99.7% recovery rate*” (Facebook); “*like the 99.8 survival rate already without a vaccine*” (Facebook). This negatively impacted perceptions of the importance of twice-weekly testing: “*8 tests a month for everyone for a virus 99.7% don’t need to worry about….*” (Facebook).

Additionally, some individuals perceived COVID-19 would not be severe due to their own natural immunity: “*Our immune systems are made to fight anything that makes us ill and antibodies will form to protect us whenever needed*” (Facebook). For some, the importance they placed in natural immunity had a negative impact on their perceptions of twice-weekly testing: “*we’re being told that perfectly healthy people need weekly tests and a vaccine for a virus their immune system can already cope with”* (Twitter).

#### Low perceived risk of contracting COVID-19

There was a lack of engagement in the twice-weekly tests when individuals felt they had a low risk of contracting COVID-19. One reason people believed they were at low risk of contracting COVID-19 was that they had limited contact with others: “*there’s no point in using it as I’ve not been outside the house since it arrived*” (News Article). Another reason was that some felt that, having worked throughout the pandemic, they had not needed a test before and did not need one now: “*I have never had a test and been working away thro this bs n will never take the test*” (Facebook); “*Plus I worked all through and never needed one then*” (News Article).

#### Low perceived severity after vaccination

Individuals also perceived COVID-19 to not be severe after vaccination. People reported the vaccine will reduce transmission: “*Infection rates will fall because the vaccine hugely reduces the risk of you transmitting the virus*” (Facebook), and offer more protection than tests: “*People have tested clear and 12 hours later got full COVID. Vaccines are the only way to protect those around you*” (Facebook). This led to perceptions that the tests were pointless in comparison to the vaccination programme: “*I don’t see the point in these tests given our heavy vaccination programme*” (News Articles).

It was also perceived that once individuals were vaccinated tests are not necessary: “*Well surely if you’ve had both if your vaccines you shouldn’t need to get tested*” (Facebook). It was thought that having the vaccine would mean individuals were well protected from COVID-19 and thus do not need to take a test: “*Surely once vaccinated we are out of the danger zone for both giving and receiving of bugs!*” (Facebook).

A consequence of the twice-weekly testing policy was that it led individuals to question the efficacy of the vaccine: “*well whats point of having vaccine then*” (Facebook); “*Also, if your vaccines work, can you kindly explain the point of this?*” (Twitter).

### Mistrust in the Government

There was widespread mistrust in government that related to: lack of government ability to implement an effective testing programme; lack of faith in government handling of the pandemic; and ulterior motives for introducing mass testing.

#### Lack of government ability to implement an effective testing programme

Individuals felt that the Government were late to mass testing and should have implemented this scale of testing sooner in the pandemic: “*which should have been implemented 12 months ago. Instead they stopped community testing, which let the virus spread rampantly during the summer*.” (Facebook); and also, that they would not be able to source enough twice weekly tests for the population due to them not being able to source enough tests in the past.

> “*Soooo they are going to magically be able to test 68mil [people]…Twice a week now???? Am I wrong in thinking they haven’t even come close to been able to test that much for the past year let alone twice a week*.” (Facebook)

#### Lack of faith in government handling of the pandemic

There was widespread mistrust in the Government’s handling of the pandemic, including perceptions that the Government had underestimated the severity of the virus, had not acted quickly enough, and had not enforced public health interventions strongly enough: “*I blame the government for a great deal in this. Too much was left too long and the laws on restrictions were too easily ignored*” (Facebook); “*So the fault is in the government downplaying the virus at the start*” (Facebook).

This mistrust in the handling of the pandemic was a barrier to engaging in the twice weekly tests as well as other preventive behaviours: “*The government can stick their tests, masks,track and trace,jabs and jab passports where the sun doesn’t shine […] The damage they have coursed [sic] is criminal and however long it takes they will be held accountable for their crimes*.” (News Articles)

#### Ulterior motives for introducing mass testing

There was a perception that there were ulterior motives for bringing in regular testing, with several ulterior motives being suggested. First, it was thought the testing programme was being introduced to justify the need to extend lockdown or enforce a lockdown at a later date: “*Is this some kind of agenda to then enforce another lockdown at a later date?!”* (Facebook); *“Just the false positives of them will permit the government to shut you all back down again”* (Facebook).

Second, it was perceived that the testing programme was being used as a way to control the population and suppress freedom: “*this is just another way of control no way*” (Facebook); “*More control and people just don’t see it. What next will they be ordering us to do? I’m beginning to feel like a leeming*” (Facebook).

Third, it was suggested that the tests were being introduced to create fear surrounding COVID-19: “*they are there to create yet more fear*” (Facebook), and stopping testing would reduce the fear of COVID-19: “*No testing = No more panic*” (Twitter).

Fourth, it was perceived the tests were being rolled out for financial benefit: “*All going to line the pockets of tory party funders and we, the people are paying through our taxes*” (Facebook). This feeling of mistrust was a reason why some individuals did not want to take a test: “*Someone’s getting rich off of all these pcr tests. I won’t be taking part*” (Facebook).

### Concern about taking a test

Another barrier to engaging in testing was concern about taking a lateral flow test. These concerns included: perception that twice-weekly tests are not normal, concern over what tests are used for, discomfort associated with testing, and perceived health risks from testing.

#### Perception that twice-weekly tests are not normal

Some suggested that it was not normal to conduct tests twice a week: “*Having to take a test to leave your house is not normal life*” (Facebook); “*How is taking 2 tests a week normal*” (Facebook). Additionally, there were concerns over how long the testing policy would continue for: “*For how long, 1 week, 1 month, 1 year? Or for ever, if its for ever no thanks*” (Facebook).

#### Concern over what tests are used for

Some were concerned that the tests may be used to collect DNA: ***“****How many times do you want a sample of my dna????*” (Facebook); and these samples may be used to make people ill: “*it’s a great way to get peoples DNA and to make them even sicker*” (Facebook).

Additionally, there were also concerns over the collection of personal information such as phone numbers, which acted as a barrier to taking a test: “*I’ve been getting texts, calls and spam emails since registering negative Covid tests. Won’t be doing that again*” (Facebook); “*when you report the findings, the government get loads of useful data about you, including your phone number*” (Facebook).

#### Discomfort associated with taking the test

Another concern was the potential discomfort associated with getting a test: “*IF you fancy having your nose prodded and your throat twice a week go ahead but not me my nose and throat wouldn’t stand for it*” (Facebook). Individuals also noted that swabbing their throat and nose for a test made them ill: “*when I did do a test it made me violently sick from gagging, don’t wanna go thru that again unless I have to!*” (Facebook); “*I wont be taking any more test have had two and still have nose bleed*” (Facebook), or that they have a medical issue which makes getting a test more painful: “*I have a deviated septum and a hole in-between my nostrils that blocks off one of my nostrils and causes sinus issues*” (Facebook).

#### Perceived health risks

Some were concerned that the tests were dangerous, with concerns largely revolving around the swabs being sterilised in ethylene oxide: “*they are giving everyone cancer as they are dipped in ethylene oxide*” (Facebook); “*Just think of all that Oxide going into your body up your nose into your brain… They must love it…. death death death*” (Facebook). This led to the perception that the ethylene oxide would cause cancer: “*The test have a carcinogenic ethanol oxide on …. causes cancer….. think about it whats the biggest killer with no cure?”* (Facebook), and resulted in people not being willing to take a test: “*it’s a no thanks for me I don’t want cancer from it*” (Facebook); “*They’re coated in ethylene oxide, a proven carcinogenic, so no I will not be taking these tests*” (Twitter).

### Perceived ineffectiveness of testing policy

Some highlighted concerns about the effectiveness of the testing policy for controlling COVID-19, with concerns including: perceived inaccuracy of the tests; potential negative impact of a negative result; perception that asymptomatic individuals are not infectious; perception that uptake of tests will be too low; and lack of trust in others to test honestly.

#### Perceived inaccuracy of the test

Some individuals believed that the tests are not accurate: “*The tests that have been proven not to work*” (Twitter); “*these tests are highly unreliable*” (Facebook). Tests were believed to be inaccurate due to giving false negatives: “*they produce a lot of false negatives*” (Facebook) and false positives: “*Cases have been proven 94% false positives*” (Twitter); “*What’s the point in a test that can give up to 50% false positives?*” (News Article). This concern over the accuracy of the tests had a negative impact on intended engagement with twice-weekly testing: “*test twice a week with a test that they have admitted is still giving false positives as well as false negatives […] SCAAP THE TESTING IT’S USELESS*” (Facebook); “*How on earth is an unreliable test going to work? The only good place for these joke of a ’test’ is on a bonfire!!*” (Facebook).

#### Potential negative impact of a negative result

Participants also believed that tests were not useful as a negative test result only reflected your status at the time of testing: “*remember that a negative test doesn’t mean you’re uninfected*” (Facebook); “*A negative test proves nothing except a person probably wasn’t infected at the time they took that particular test*” (Twitter). In addition, some were concerned that a negative test result may influence behaviour and lead to individuals not adhering to other preventive health measures.

> “*The problem is that if you are told you are ‘negative’ you are more likely to become overconfident and relax your precautions. I am not against the LFT, what I want is for people to be told what the results mean, and in particular that a ‘negative’ result doesn’t mean you are not carrying the virus*” (Facebook).

This then led to participants to not want to engage in testing: “*you can test negative one day and positive the very next […] I will not succumb to testing*” (Facebook).

#### Perception that asymptomatic individuals will not be infectious

There was the perception that asymptomatic individuals did not need a test as they were not ill: “*if you need a test to tell you you’ve got covid then you are very clearly not I’ll enough to die from it, even if you are you’ve already got it so it’s a bit late*” (Facebook); “*Why do i need to test myself, when theres nothing wrong with me?*” (Twitter). However, some individuals did state they would get a test if they were ill: *“I will never take a test unless I have symptoms”* (Facebook).

Some individuals also perceived that testing asymptomatic people was not effective as they could not spread COVID-19: “*asymptomatic people rarely spread any coronavirus,there’s plenty of papers around proving it…once those at risk are injected the rest of don’t need it*” (Facebook); “*asymptotic spread is extremely minimal*” (Facebook).

### Perception that uptake of tests will be too low

There was a perception that engagement in testing would be low: “*I doubt many people will take the tests*” (News Article). One reason uptake was perceived to be low was due to the lack of engagement with other COVID-19 preventive behaviours: “*too many refuse to do either [wear a mask and get vaccinated], without medical reasons. Do you think those people are going to self-test twice a week?*” (Facebook), as well as the concern over the accessibility of these tests that may limit engagement: “*There will be a lot of elderly people who won’t be able to do this*” (Facebook). Some people also linked the perception that others are not engaging in testing to their own lack of engagement with testing: “*Not all adults.. There will be plenty that won’t have this.. Just like the jab.. I won’t be doing either..”* (Facebook).

#### Lack of trust that others will test honestly

There was also scepticism over whether people would engage with the tests honestly, with some suggesting that people may lie about their results or not report the result of the test on the Government website: “*Do you need to do it at home and just say what the results are? So you can say its positive or negative and just lie about it*” (Facebook); “*a lot of people won’t register them (their test result)”* (Facebook). It was also suggested that people may lie about their result to avoid self-isolation or because they did not want to miss out on social events: “*This will not work because you’re relaying on people to be 100% honest with their results. What happens if someone gets a positive result, but feels fine in themselves because 1 in 3 people are asymptotic, they won’t self isolate or stay off work*” (Facebook).

### Perceived negative impact of twice-weekly testing policy

As well as concerns over the perceived lack of effectiveness of the twice-weekly testing policy, some highlighted potential negative impacts of the policy, including: the inappropriate use of public money; the financial impact of self-isolation; and the environmental impact.

#### Inappropriate use of public money

It was suggested that the testing initiative was not an appropriate use of public money: “*its a huge waste of money and completely pointless”* (Facebook); “*What a waste of public funds*” (Facebook), and that they were a waste of tax payers money: “*It’s all been a waste of tax payers money*” (Facebook); “*Just a total waste of more taxpayer billions. I certainly won’t be taking part in this testing*” (News Articles).

Relatedly, there was also concern about the impact of the testing programme on resources for other parts of the healthcare sector: “*With the money they are wasting on covid culture we could have GP surgeries that welcome patients, hospital that are free from infection, minor but life changing ops carried out within a few weeks*” (Facebook); “*Rather the money went into ploughing through the waiting lists of non Covid patients*” (News Article). There was also a concern over a lack of focus on rescheduling postponed surgeries, seeing a GP, and the rise in other diseases: “*Remember that there is also a huge backlog of elective surgeries and treatments that were postponed*” (Facebook). It also linked with the sub-theme of asymptomatic illness; people did not understand why resources were directed into testing asymptomatic people instead of people with other illnesses.

> “*Why do i need to test myself, when theres nothing wrong with me? Theres millions of genuinely I’ll people in the UK who cant see a GP or get proper treatment for serious illness.. I’d say the costs and resources to continuously test non- Ill people should be used elsewhere….*” (Facebook).

#### Financial impact of self-isolation

Analysis highlighted that the lack of financial support for people self-isolating was also a key barrier to engaging in the testing programme: “*isolation pay should be implemented or it will never go away*” (Facebook). This led to a suggestion that people would not want to engage in twice weekly testing because they could not afford to self-isolate: “*All the urging in the world won’t persuade any who can’t afford to isolate if test is positive*” (Facebook); “*many people cant afford to test, a positive test mean no work, no money*” (Facebook).

#### Environmental impact of tests

Other comments revolved around concerns about the negative environmental impact of the tests: “*The environmental impact bothers me – the whole kit goes straight in the bin*” (Facebook); “*What about all the pollution from all this discarded swabs, ppe etc*” (News Article). This led to some individuals not wanting to get tested due to the amount of waste the tests will produce.

> “*Each box consists of hundreds of pieces of plastic. If millions of us have these kits, that’s going to be billions of pieces of plastic […] We only have a few years in which to prevent complete climate breakdown and we’re doing the opposite of what we should be doing. I’ve declined another box of tests*.” (Facebook)

### Wanting to protecting others

Wanting to protect others was a key facilitator of engagement with the testing policy. This included wanting to protect friends and family: “*I can keep my family and friends safe knowing if I’m clear of the virus*” (Facebook); and preventing further outbreaks “*I see it’s a duty of care for everyone to be doing it to stop new outbreaks*” (Facebook). Individuals reported feeling safe meeting others after they had done their test: “*people want to do them for their own peace of mind and it allows them to feel safe meeting their family*.” (Facebook). There was also a perception that those that are not engaging in the testing may end up being responsible for transmitting the virus: “*those that choose not to and go around spreading it can be responsible for someones death without even knowing it*” (Facebook).

There was also a perception amongst some that the vaccine is not effective at reducing transmission: *“you can still catch and spread it even after having the vaccine*” (Twitter), which highlighted the importance of engaging in the twice-weekly testing even when fully vaccinated.

> “*u can still carry virus with the jab just not get as poorly but then pass it on to a non vaxer who could end up in the hospital I think its a good idea.”* (Twitter)

### Positive perceptions of tests

Some expressed positive perceptions of the tests, including that the tests are accurate, accessible, and quick to do.

#### Tests are accurate

Some people were in favour of the policy because they believed the tests were accurate: “*The probability of a false positive in the lft is less than 1 in 1000*” (Facebook). In most instances the positive attitude towards the accuracy of tests came from previous experiences of testing: “*i was told at my local testing sight you cannot get a false negative with the quick tests*” (Facebook); *“Our school has done thousands of tests, no false positives*” (Facebook). It was also highlighted that the tests were accurate for largescale use and to identify outbreaks: “*It is perfectly fine for it’s use as a large scale screening test”* (Facebook).

#### Tests are accessible

Positive perceptions also included tests being widely available: “*they are easily available*” (Facebook); “*theyre going to be available from your GP’s surgery, chemists and covid testing centres as well as being able to order them online, can’t get more widely available than that*” (Facebook).

#### Tests are quick

Individuals noted that tests were quick to complete: “*Do 2x simple tests and we can all move on. Zero harm, 5mins of your time.. simple*” (Facebook); “*The test takes less than a minute*” (Facebook), and that the results were quick: “*asymptomatic testing is well easy and results usually in an hour*” (Facebook).

### Desire to return to normal

Another reason that individuals gave for wanting to engage in the testing was the anticipation of returning to ‘normal’: “*I’m all for it if it means getting back to normal*” (Facebook), including restrictions being eased: “*It’s a small price to pay to be able to ease restrictions further and be able to travel and see family*” (Facebook), and reduced likelihood of further lockdowns: “*This minimises chances of another lockdown*” (Twitter).

### Perceived efficacy for reducing asymptomatic transmission

The final reason that people gave for engaging in tests was to reduce asymptomatic transmission. Individuals highlighted the importance of twice-weekly testing due to asymptomatic transmission: “*the purpose of testing ’perfectly healthy people’ is finding asymptomatic carriers, you know, to stop them spreading it without knowing*” (Facebook). Others compared the importance of asymptomatic testing to screening for other diseases such as cancer: “*Really? Have you never heard of HIV tests, cancer screening*” (Facebook); “*shall we stop testing for cancer while we’re at it? Can adjust those cancer statistics by just not being diagnosed right?*” (Facebook).

## Discussion

In this study we sampled social media comments relating to the twice-weekly testing policy in England to identify attitudes towards the policy, as well as barriers and facilitators to engaging in twice-weekly testing. Attitudes towards twice-weekly testing were predominately negative, with most comments reflecting barriers rather than facilitators of testing. Whilst some individuals highlighted perceived benefits of twice weekly testing, most people did not perceive testing to be an effective way out of the pandemic and described a range of barriers that would discourage them from engaging with twice weekly testing.

### Barriers to testing

Barriers to engaging in twice-weekly testing included low perceived risk of COVID-19, mistrust in authorities, concern about taking a test, perceived ineffectiveness of testing policy, perceived negative impact of twice-weekly testing policy, and perceived efficacy of the vaccine.

Individuals perceived a low risk from COVID-19, both in terms of severity of the virus and the likelihood of contracting it. This low perceived risk from COVID-19 led to a belief that testing was unnecessary, and therefore reduced intentions to engage with twice weekly testing. This is in line with previous research showing that individuals who perceive COVID-19 to be less of a risk engage in less preventative behaviour [11, 12]. Additionally, the vaccine rollout reduced the perceptions of the severity of COVID-19, individuals preferred vaccinations as a route of the pandemic, rather than testing, with some suggesting they or others did not need to engage in the testing as they had been fully vaccinated.

Mistrust in the Government was an additional barrier to engaging in testing. This mistrust included a lack of confidence in the Government’s ability to conduct mass twice-weekly testing and handle the pandemic, as well as mistrust about the purpose of the twice weekly testing policy (e.g. to induce fear, exert control, or justify further lockdowns) and the motives of policy makers (e.g. financial gain). Research has shown that a lack of confidence and mistrust in the Government is a barrier to engaging with asymptomatic testing [6] and leads to less engagement in COVID-19 protective behaviours [13, 14, 15]. Uncertainty around the purpose of testing was identified as a barrier to getting tested during the mass asymptomatic testing in Liverpool [6]. Communicating openly and honestly with members of the public about why certain actions are being taken has been shown to increase perceived legitimacy of authorities’ actions, and enhance adherence with recommended behaviours [16, 17]. Therefore clear, open and honest communication about why the twice-weekly testing initiative has been rolled out may reduce mistrust in the motivation behind twice weekly testing and improve testing uptake.

Another barrier was concern about taking a lateral flow test, which included the concerns that tests were being used to collect personal data, the potential health risks from the tests, discomfort of testing, and perception that taking a test twice a week was not normal and concerns over how long this would last. This is in-line with previous research that has identified concerns about use of personal data and discomfort of swabbing as barriers to testing [7, 18].

The effectiveness of testing to help control the pandemic was also questioned, with a particular concern being that the tests are not accurate, and that false negatives and false positives are common. Perceived inaccuracy of tests has been shown to be an important factor in determining uptake of testing [18]. Additionally, there were concerns that a negative test result only provides a snapshot of an individual’s likelihood of transmitting the virus and that others will not test honestly, both of which impacted reported engagement with twice weekly testing. The test was also perceived to be unnecessary due to a belief that asymptomatic individuals will not spread the virus.

In addition to concerns about the ineffectiveness of the testing policy, individuals also highlighted some potential negative impacts of the policy. These included the negative environmental impact of so many tests, a perception that the twice weekly testing policy was a waste of public money, and the negative financial impact on those who need to self-isolate. Lack of support for those self-isolating has also been identified as a barrier to testing in previous research [6, 7], and therefore financial support should be provided to everyone who needs to self-isolate, in order to encourage uptake of testing [19].

### Facilitators of testing

Whilst most comments related to barriers to testing, some people also talked about reasons that they would engage with twice weekly testing. Common reasons included wanting to protect others and wanting to return to normality; this is in line with previous research into barriers and facilitators to engaging with asymptomatic testing in Liverpool [6]. Some people also felt that the tests were accurate, accessible, and provided rapid results; this was often based on previous experiences of taking a test. This finding supports previous research that has identified speed and convenience of testing as important facilitators of testing uptake [7, 8]. In this aspect the twice-weekly testing policy has a clear advantage over previous mass asymptomatic testing, as it enables people to take a test at home and get the results in under 30 minutes [1], rather than having to go to a testing site [20].

### Recommendations

We recommend, based on the findings, that to increase engagement with the twice-weekly testing authorities should: 1) communicate openly and honestly about the purpose of introducing twice-weekly testing, including the reasoning behind two tests a week and how long this policy is intended to last; 2) provide information on the efficacy of using tests to help control the pandemic, including the accuracy of the tests and the role in reducing asymptomatic spread; 3) provide financial support for those that are required to self-isolate; communicate the purpose of testing for fully-vaccinated individuals, without undermining the role of vaccines; 5) continue to make tests free and easily accessible via free delivery to homes or widely available to collect.

### Limitations

A key limitation of this study is that the results may not be representative of all people in England due to sampling social media comments [21]. There are demographic differences between those who use social media and those who do not [22], and therefore findings may not be representative of those who do not use social media. Additionally, the views of individuals who comment may be skewed towards more extreme perspectives and thus not representative of all attitudes towards twice-weekly testing. Despite these limitations, social media data does provide real-time data of public health behaviour [23].

## Conclusions

To conclude, the results show several barriers to engaging with twice weekly testing, as well as some facilitators. Barriers to engaging in twice-weekly testing include low perceptions of risk from COVID-19, mistrust in the government, and concern about taking a test. There was also a perception that the twice weekly testing policy would not be effective or would have a negative impact. These barriers all negatively impacted willingness to engage with the twice weekly testing policy. Facilitators to engaging in testing included wanting to protect others, wanting to return to normal, and a perception that tests are accessible and accurate. Based on these findings, several recommendations can be made to increase engagement with twice weekly testing: communicate openly and honestly about the purpose of testing; provide information about the accuracy of tests; provide financial support for those required to self-isolate; and emphasise accessibility of testing.

## Data Availability

The data used in the current study are available from the corresponding author on reasonable request.

## Declarations

### Ethics Approval and Consent to Participate

Public Health England (PHE) Research Ethics and Governance Group (REGG) exempted this study from requiring ethical approval because the study used publicly available data.

### Consent to Participate

Not applicable as no individuals participated in the study and no identifiable information was included.

### Competing Interests

No competing interests.

### Funding

This research received no specific grant from any funding agency in the public, commercial or not-for-profit sectors.

### Author Contributions

All authors contributed to the design of the study. AD sampled the data and conducted the data analysis. AD wrote the paper with CR and HC commenting, reading, and approving the final manuscript.

## Acknowledgments

We thank Lyndsay McAteer for assistance with data analysis.

